# Unveiling the Best Asthma Control Regiment: A Comprehensive Meta-Analysis and Meta-Regression of Efficacy, Safety, and Cost-Effectiveness

**DOI:** 10.1101/2025.03.09.25323633

**Authors:** Kevin Christian Tjandra, Arlina Dewi, Fahrul Nurkolis

## Abstract

**Background:** Asthma remains a major global health concern, requiring effective and cost-efficient treatment strategies. Long-acting beta-agonists (LABAs), particularly in combination with inhaled corticosteroids (ICS), are widely used to manage symptoms and prevent exacerbations. However, uncertainty persists regarding the optimal regimen in terms of effectiveness, safety, and affordability, especially in resource-limited settings. This study evaluates the effectiveness, safety, and cost-efficiency of ICS/LABA regimens versus alternative asthma treatments, focusing on exacerbations, lung function, symptom control, and cost-effectiveness, especially in resource-limited settings.

**Methodology:** An extensive search across databases such as Scopus, PubMed, Cochrane, and others was conducted, focusing on asthma treatments. Our inclusion criteria covered RCTs on asthma, ICS/LABA regimens, comparisons with alternative treatments. Key outcomes included incidence of exacerbation, Forced Expiratory Volume (FEV1), Peak Expiratory Flow Rate (PEFR), Asthma Control Questionnaire (ACQ-5) scores, and cost-effectiveness, covering a broad range of patient demographics. The analysis compared ICS/LABA combinations to alternative therapies, synthesizing outcomes using standardized mean differences (SMD) and odds ratios (OR) with 95% confidence intervals (CI) through random-effect models. Quality assessment followed the Cochrane Collaboration’s RoB-2 tool, and meta-analyses were performed in R-Studio. The cost analysis was also conducted utilizing tornado plots, which outline what influences costs.

**Results:** A total of 5,083 studies were identified, with 11 meeting inclusion criteria (19,905 participants). Meta-analysis showed ICS/LABA combinations significantly reduced exacerbations (RR = 0.67, 95% CI: 0.54–0.84) but had minimal impact on FEV1 and ACQ-5 scores. PEFR improved significantly (SMD = 1.25, 95% CI: 1.09–1.40). Meta-regression indicated that age and follow-up duration had minor effects on PEFR but did not influence exacerbations. Cost analysis found ICS/LABA treatments cost up to $150, while terbutaline costs under $50, offering up to 66.7% savings. Further research is needed to balance cost-effectiveness with treatment efficacy in asthma management.

**Conclusion:** This study confirms that ICS/LABA combinations reduce exacerbations and improve PEFR, though with minimal effects on FEV1 and ACQ-5 scores. Meta-regression showed slight PEFR variations by age and follow-up. Cost analysis suggests up to 66.7% savings with terbutaline, though ICS/LABA offers superior outcomes. Balancing efficacy and cost is crucial for optimizing asthma management.

## Introduction

Asthma remains a significant global health burden, affecting millions of individuals and contributing to high rates of morbidity, healthcare utilization, and economic costs. Effective management is critical to preventing exacerbations and improving patients’ quality of life. Among the available treatments, LABAs, particularly in ICS, are widely used to control symptoms and reduce the frequency of asthma attacks. This combination provides sustained bronchodilation and anti-inflammatory effects, making it a cornerstone of asthma therapy. However, despite its widespread use, uncertainties remain regarding the optimal LABA regimen in terms of efficacy, safety, and cost-effectiveness, particularly in resource-limited healthcare settings[1,2].

Several studies have highlighted the benefits of ICS/LABA combinations in reducing asthma exacerbations and improving respiratory function. Beasley et al. (2022) demonstrated that LABA-ICS therapy significantly lowers exacerbation rates in patients with persistent symptoms, while Sobieraj et al. (2018) reported improved symptom control and enhanced quality of life. Conversely, safety concerns persist, as Salpeter et al. (2006) found that using LABAs without ICS may increase the risk of severe asthma-related complications. These findings underscore the importance of selecting appropriate treatment strategies to maximize benefits while minimizing risks[1–3].

Beyond clinical outcomes, cost remains a key factor influencing asthma treatment choices, particularly in primary care settings where financial constraints impact decision-making. While LABA therapy may involve higher initial costs, Fingelton et al. (2017) found that ICS/LABA combinations ultimately reduce long-term healthcare expenses by decreasing emergency visits and hospitalizations. Additionally, Almutairi et al. (2023) estimated that integrating LABAs into structured asthma management programs could lower overall treatment costs by up to 30%, making cost-effectiveness a crucial consideration in treatment selection[4,5].

Despite the growing body of research on asthma management, previous systematic reviews and meta-analyses have focused primarily on either clinical efficacy or safety outcomes, with limited emphasis on cost-effectiveness. Many existing reviews lack a comprehensive evaluation that integrates effectiveness, safety, and financial implications into a single assessment. Additionally, prior studies have not extensively explored how key patient factors—such as age, gender distribution, and follow-up duration—affect treatment outcomes through meta-regression analysis. By addressing these gaps, this study provides a more holistic and data-driven evaluation of ICS/LABA therapies, helping to guide both clinical and economic decision-making.

To fill these gaps, this study aims to systematically evaluate the effectiveness, safety, and cost-efficiency of ICS/LABA regimens compared to alternative asthma treatments, particularly in resource-limited settings. The analysis incorporates clinical outcomes, such as exacerbation rates, lung function (FEV1, PEFR), symptom control (ACQ-5), and cost-effectiveness, to provide comprehensive insights into optimal asthma management strategies.

## Methods

### Eligibility criteria

This systematic review followed the Preferred Reporting Items for Systematic Reviews and Meta-Analyses (PRISMA) guidelines, ensuring a transparent and reproducible approach [6]. We included original research published between 2013 and 2024, with the final literature search conducted on October 20, 2024. Eligible studies were randomized controlled trials (RCTs) that enrolled asthma patients across diverse age groups, sexes, and geographical regions. The primary intervention of interest was inhaled corticosteroids (ICS) combined with long-acting beta-agonists (LABA), administered in any regimen. Comparators included alternative asthma treatments, such as terbutaline, montelukast, fluticasone-salmeterol, or placebo. Eligible studies were required to report at least one relevant outcome, including exacerbation incidence (moderate or severe), FEV₁, PEFR, ACQ-5 scores, or cost-effectiveness.

We excluded non-comparative studies, technical reports, editorials, narrative reviews, systematic reviews, meta-analyses, in silico, in vitro, and in vivo studies, as well as conference abstracts, scientific posters, and research proposals. We also excluded studies not written in English, studies with incomplete data, and those that were unrelated to asthma treatment or pharmacotherapy. These exclusions were necessary to maintain a focus on clinically relevant evidence and avoid introducing indirectness or reporting bias into the analysis. The potential impact of these exclusions, including language bias and the exclusion of gray literature, is further addressed in the Limitations section.

To enhance reliability and minimize selection bias, study selection was conducted through a double-review process. Two independent reviewers screened titles and abstracts, followed by a full-text review for eligibility. Any disagreements between reviewers were resolved through discussion, and if consensus could not be reached, a third senior reviewer provided the final decision. This multi-step review process, combined with predefined inclusion and exclusion criteria, ensured that the selection process was both systematic and reproducible. Furthermore, studies included in this review adhered to the PICO framework, where the population consisted of asthma patients, the intervention included ICS/LABA combinations (such as budesonide– formoterol or beclomethasone–formoterol), the comparator involved placebo, terbutaline, montelukast, or other asthma treatments, and outcomes included exacerbation incidence, FEV₁, PEFR, ACQ-5 scores, and cost-effectiveness, with detailed extraction of sample size, follow-up duration, gender distribution, and mean participant age.

### Data search and selection

The literature search was conducted using a comprehensive multi-database strategy across Scopus, PubMed, ProQuest, ScienceDirect, Sage Pub, and Cochrane, ensuring broad coverage of relevant literature. Search terms were developed using Medical Subject Headings (MeSH) and free-text keywords, combined using Boolean operators to balance sensitivity and specificity. The final search string applied across databases was:

(‘Asthma’) AND (‘Exacerbation’ OR ’FEV1’ OR ’PEFR’ OR ’ACQ-5 Score’) AND (‘Budesonide’ OR ’Formoterol’ OR ’Beclomethasone’ OR ’ICS/LABA’ OR ’Terbutaline’ OR ’Montelukast’).

The search covered publications from database inception through October 20, 2024, with a particular focus on studies published in the past 10 years to capture current clinical evidence. Filters were applied to restrict results to English-language, peer-reviewed original research articles. To manage references and facilitate collaboration across the research team, we used Mendeley Group Reference Manager, enabling centralized access, screening, and annotation. After removing duplicates, five authors (KCT, FN, HANA, AD) independently screened the titles and abstracts, followed by a full-text eligibility review conducted by two reviewers (KCT and FN). Any discrepancies were resolved through discussion, and AD provided the final decision when consensus was not achieved. This multi-step process ensured that study selection was transparent, reproducible, and minimized potential selection bias.

### Study Risk of Bias Assessment (Qualitative Synthesis)

The risk of bias assessment was conducted independently by five authors using validated tools. Since all included studies were randomized controlled trials (RCTs), we applied the Cochrane Collaboration’s Risk of Bias version 2 (RoB-2) tool, which systematically evaluates five key domains: (a) the randomization process, (b) deviations from intended interventions, (c) missing outcome data, (d) measurement of outcomes, and (e) selection of reported results. Each study was assigned a classification of low risk, some concerns, or high risk of bias for each domain. Any disagreements between authors during the assessment process were resolved through group discussion. If consensus could not be reached, a senior author (AD) provided the final decision.

To assess the certainty of evidence across outcomes, we employed the Grading of Recommendations, Assessment, Development, and Evaluations (GRADE) approach. This framework considers five factors that can influence the certainty of evidence: risk of bias, inconsistency (heterogeneity), indirectness, imprecision, and publication bias. Based on these criteria, each outcome was ultimately rated as having high, moderate, low, or very low certainty. By combining the RoB-2 assessment for individual study quality with the GRADE framework for overall evidence certainty, this process ensures a transparent and standardized evaluation of both study validity and the strength of the evidence as a whole.

### Data extraction

After completing the final screening process, relevant data from the selected studies were systematically extracted and organized into a Google Spreadsheet. The data table encompassed essential study characteristics, including the author, year of publication, country, study design, sample size, mean age, gender distribution, type of intervention, type of control, follow-up duration, and primary outcomes. Key outcomes of interest included incidence of exacerbation, FEV1, PEFR, ACQ-5 scores, and cost-effectiveness.

The data extraction process was conducted independently by three authors (KCT, FN, and HANA) to ensure accuracy and consistency. For each outcome, detailed numerical data such as means, standard deviations (SDs), and additional relevant metrics were recorded. Any discrepancies encountered during the extraction process were resolved through discussion among the authors, and in cases where a consensus could not be reached, AD made the final decision. This approach ensured a rigorous and reliable compilation of data for subsequent analysis.

### Statistical Analysis

All statistical analyses were conducted using R Studio to ensure precision and reliability in interpreting the results. Given the anticipated variability in study populations, interventions, and follow-up durations, we applied a random-effects model for all meta-analyses. This model was chosen because it accounts for underlying differences across studies, making it more appropriate when dealing with heterogeneity in real-world clinical data. Since high heterogeneity was expected due to differences in study designs, populations, and interventions, the use of a random-effects model was justified. This model assumes that variations exist not only due to sampling error but also due to true differences among studies, making it suitable for our meta-analysis.

For continuous outcomes, we used the SMD with 95% confidence intervals (CI) instead of the MD. SMD was selected because the included studies reported outcomes using different measurement scales, requiring standardization for comparability. This approach ensures that variations in scoring methods across studies do not skew the overall findings. For dichotomous data, we employed the Mantel-Haenszel method for subgroup analysis to derive pooled risk ratios, as it provides a reliable estimation of effect sizes in studies with different sample sizes.

To assess heterogeneity among studies, we used the I-squared (I²) statistic and Chi-square (χ²) test. I² values were interpreted as follows: low heterogeneity (I² < 25%), moderate heterogeneity (25% ≤ I² < 75%), and high heterogeneity (I² ≥ 75%). The Chi-square (χ²) test was used to evaluate whether the observed heterogeneity was statistically significant. Since substantial heterogeneity was anticipated due to differences in study populations and methodologies, I² values above 25% were considered meaningful, justifying the choice of the random-effects model. For continuous data reported as medians with interquartile ranges (IQRs) or minimum and maximum values, we applied transformation methods from Luo et al. and Wan et al.[7,8] to estimate means and standard deviations (SD). This standardization ensured consistency in data presentation and facilitated proper pooling of results.

Potential publication bias was assessed using funnel plots. If asymmetry was detected, we examined the PICO elements and outcome characteristics to determine whether the asymmetry resulted from publication bias or methodological heterogeneity. To further ensure the robustness of our results, we conducted a sensitivity analysis, restricting the meta-analysis to studies with an overall low risk of bias. This step aimed to confirm the stability and reliability of our conclusions.

To explore potential confounding factors, we performed meta-regression analysis, assessing the impact of sample size, mean age, gender distribution (male/female), and follow-up duration on exacerbation rates and PEFR. This analysis helped determine whether specific patient demographics or study characteristics influenced treatment outcomes.

In addition to clinical outcomes, we conducted a cost analysis to evaluate the economic implications of various asthma treatments. This included tornado plots to visualize cost sensitivity and identify variables that significantly impact treatment expenses, as well as cost-per-treatment calculations incorporating direct costs (e.g., medication expenses) and indirect costs (e.g., follow-up care and exacerbation management). The cost analysis was performed using R Studio, integrating financial data with clinical outcomes to provide a comprehensive assessment of treatment value. Baseline cost, lower bound cost, and upper bound cost data were sourced from The U.S. National Library of Medicine (MedlinePlus). This approach ensures that treatment recommendations consider both clinical effectiveness and financial sustainability, providing valuable insights for healthcare decision-making in asthma management.

## Result

### Study Selection

A total of 5,083 studies were initially identified across multiple databases, with the following distribution: 2,191 from Scopus, 1,151 from PubMed, 751 from Cochrane, 130 from Sage Pub, 553 from ProQuest, 180 from PLOS, 7 from EBSCO, 40 from Epistemonikos, and 80 from Springer. After filtering out records based on a 10-year publication limit and specific article types, 4,641 studies were removed, leaving 442 studies for further screening. During the screening process, 207 studies were excluded due to being unrelated to the research topic, resulting in 235 studies for full-text retrieval. All 235 studies were successfully retrieved for detailed assessment.

Of the reports assessed for eligibility, 224 were excluded for various reasons: 30 had the wrong study design, 38 did not meet the outcome criteria, 100 presented data in an incompatible format, 20 involved the wrong patient population, 15 used an incorrect intervention, 9 had the wrong comparator, and 12 addressed the wrong indication. Ultimately, 11 studies met all criteria and were included in the systematic review. This study selection process is illustrated in the PRISMA flowchart (Figure 1). The included studies were then assessed for bias using the Cochrane ROB-2 tool, with detailed results provided in the following sections of this manuscript.

**Figure 1.**
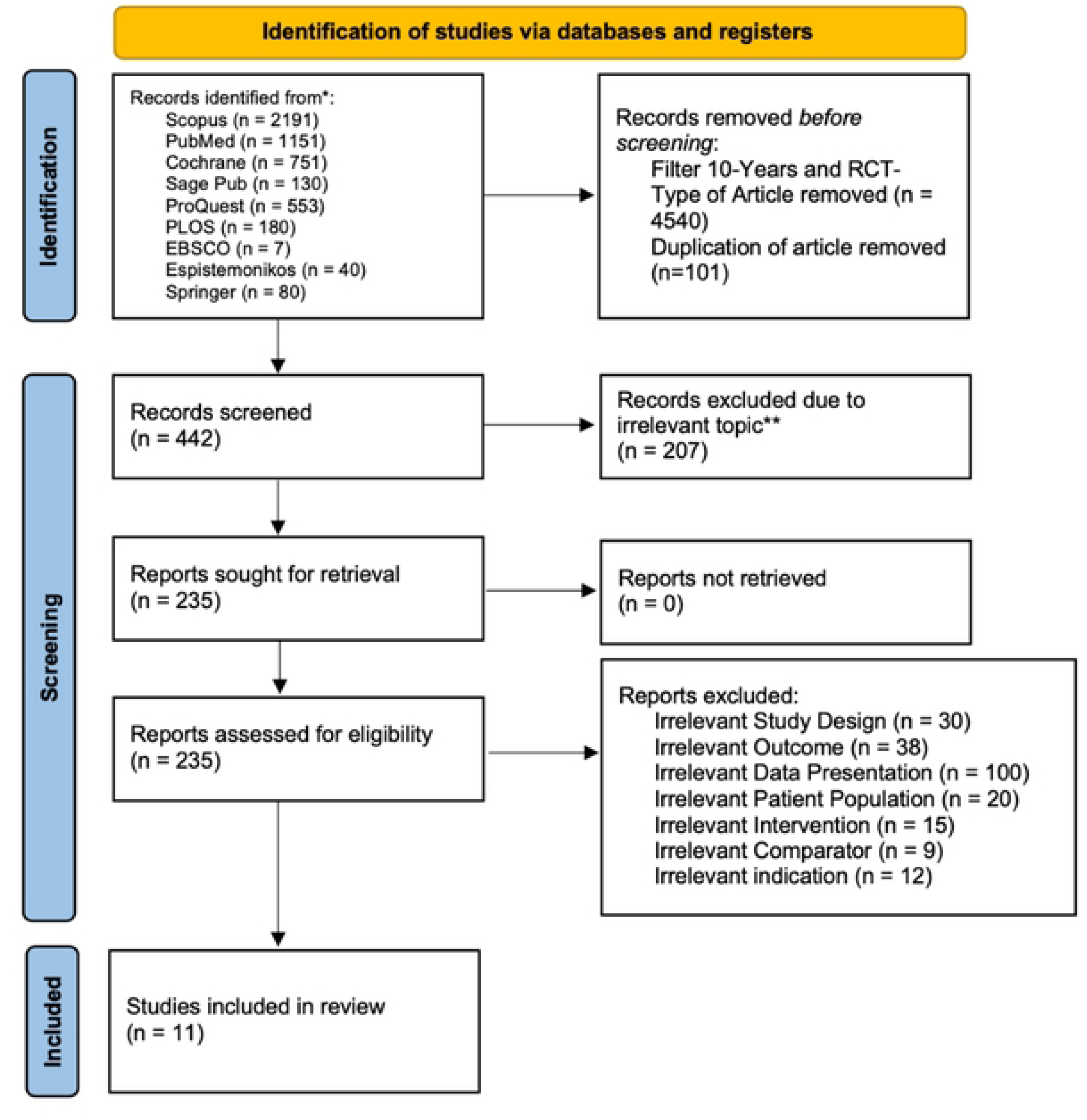
PRISMA 2020 flow diagram.

### Study Characteristics

The systematic review included 11 studies with a total of 19,905 participants, spanning across seven countries: Canada (2 studies), the United States (3 studies), China (2 studies), South Africa (1 study), Taiwan (1 study), Scotland (1 study), and Iran (1 study). These studies utilized rigorous methodologies and investigated a wide range of asthma management strategies, contributing valuable insights into treatment efficacy, safety, and cost-effectiveness.

The studies varied in design, focusing on the effectiveness of ICS and LABA in different formulations and regimens. Several studies compared budesonide–formoterol with alternative treatments such as terbutaline, fluticasone-salmeterol, and olodaterol, evaluating their impact on asthma control. These interventions included both fixed-dose and as-needed formulations, as well as maintenance therapy approaches, ensuring a comprehensive assessment of different asthma treatment strategies.

The participant demographics reflected a broad age range, with mean ages varying between 36 and 59 years, and included a balanced male-to-female distribution. Follow-up periods ranged from 1 to 12 months, allowing for the assessment of both short-term symptom control and long-term treatment outcomes.

By incorporating diverse study designs and regional populations, this review provides a robust dataset that enhances the generalizability of findings across different healthcare settings. The full study characteristics, including details on Population, Intervention, Comparison, and Outcomes (PICO), are systematically outlined in Table 1.

**Table 1.**
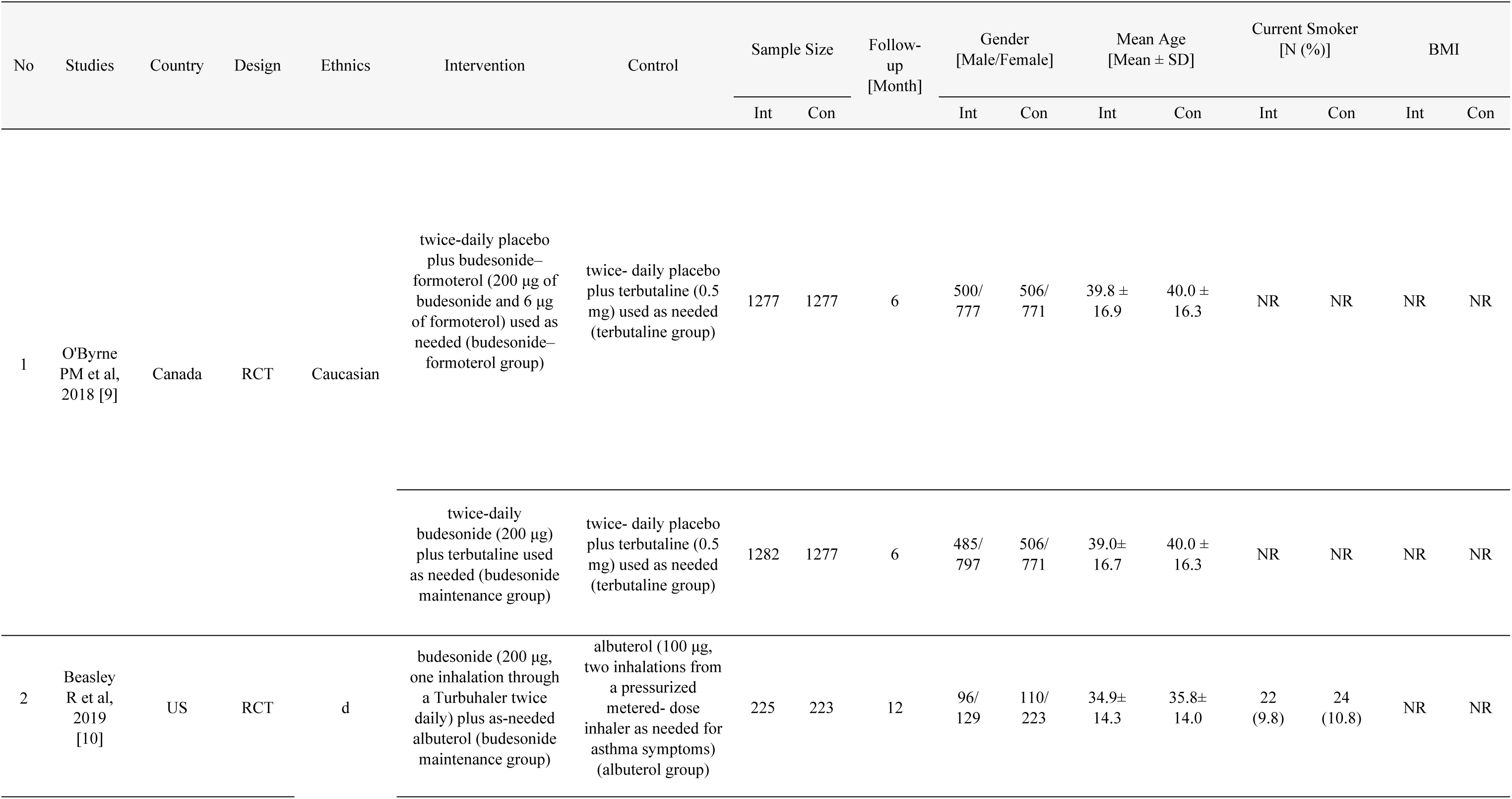
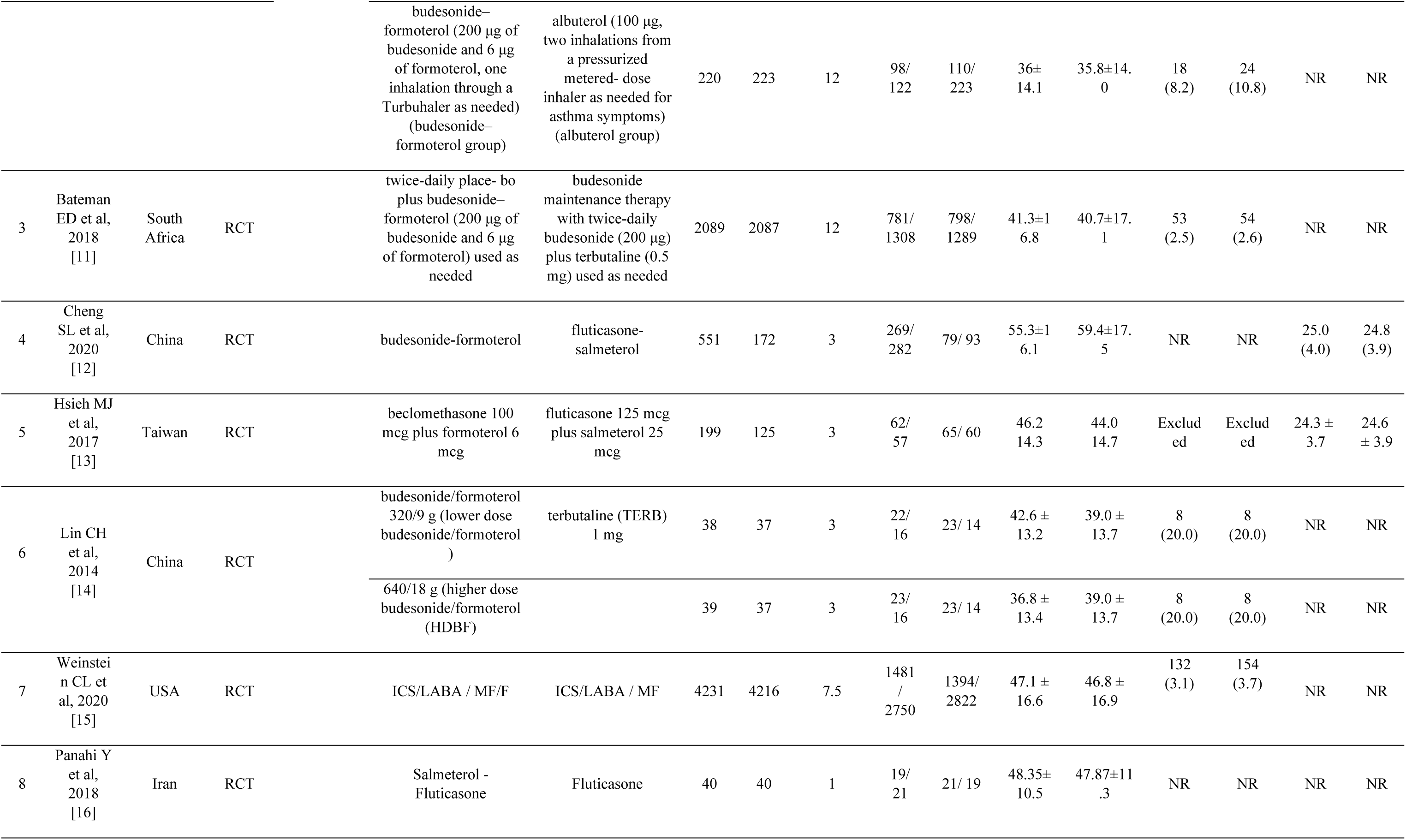
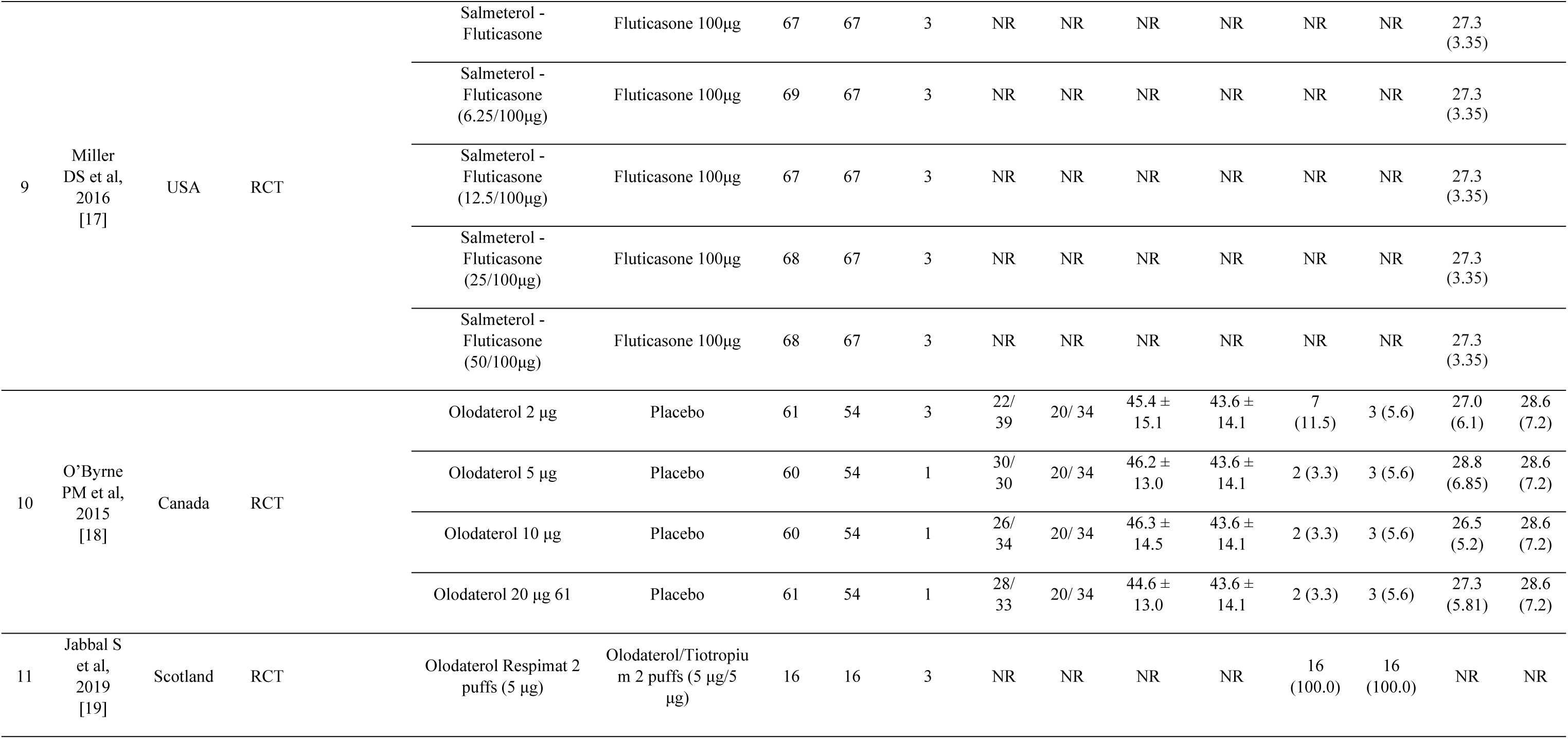
Study Characteristic.

| No | Studies | Country | Design | Ethnics | Intervention | Control | Sample Size |  | Follow-up<br>[Month] | Gender<br>[Male/Female] |  | Mean Age<br>[Mean ± SD] |  | Current Smoker<br>[N (%)] |  | BMI |  |
| --- | --- | --- | --- | --- | --- | --- | --- | --- | --- | --- | --- | --- | --- | --- | --- | --- | --- |
|  |  |  |  |  |  |  | Int | Con |  | Int | Con | Int | Con | Int | Con | Int | Con |
| 1 | O'Byrne<br>PM et al,<br>2018 [9] | Canada | RCT | Caucasian | twice-daily placebo<br>plus budesonide–<br>formoterol (200 µg of<br>budesonide and 6 µg<br>of formoterol) used as<br>needed (budesonide–<br>formoterol group) | twice- daily placebo<br>plus terbutaline (0.5<br>mg) used as needed<br>(terbutaline group) | 1277 | 1277 | 6 | 500/<br>777 | 506/<br>771 | 39.8 ±<br>16.9 | 40.0 ±<br>16.3 | NR | NR | NR | NR |
|  |  |  |  |  | twice-daily<br>budesonide (200 µg)<br>plus terbutaline used<br>as needed (budesonide<br>maintenance group) | twice- daily placebo<br>plus terbutaline (0.5<br>mg) used as needed<br>(terbutaline group) | 1282 | 1277 | 6 | 485/<br>797 | 506/<br>771 | 39.0±<br>16.7 | 40.0 ±<br>16.3 | NR | NR | NR | NR |
| 2 | Beasley<br>R et al,<br>2019<br>[10] | US | RCT | d | budesonide (200 µg,<br>one inhalation through<br>a Turbuhaler twice<br>daily) plus as-needed<br>albuterol (budesonide<br>maintenance group) | albuterol (100 µg,<br>two inhalations from<br>a pressurized<br>metered- dose<br>inhaler as needed for<br>asthma symptoms)<br>(albuterol group) | 225 | 223 | 12 | 96/<br>129 | 110/<br>223 | 34.9±<br>14.3 | 35.8±<br>14.0 | 22<br>(9.8) | 24<br>(10.8) | NR | NR |

|  |  |  |  |  |  |  |  |  |  |  |  |  |  |  |  |  |
| --- | --- | --- | --- | --- | --- | --- | --- | --- | --- | --- | --- | --- | --- | --- | --- | --- |
|  |  |  |  | budesonide–<br>formoterol (200 µg of<br>budesonide and 6 µg<br>of formoterol, one<br>inhalation through a<br>Turbuhaler as needed)<br>(budesonide–<br>formoterol group) | albuterol (100 µg,<br>two inhalations from<br>a pressurized<br>metered- dose<br>inhaler as needed for<br>asthma symptoms)<br>(albuterol group) | 220 | 223 | 12 | 98/<br>122 | 110/<br>223 | 36±<br>14.1 | 35.8±14.<br>0 | 18<br>(8.2) | 24<br>(10.8) | NR | NR |
| 3 | Bateman<br>ED et al,<br>2018<br>[11] | South<br>Africa | RCT | twice-daily place- bo<br>plus budesonide–<br>formoterol (200 µg of<br>budesonide and 6 µg<br>of formoterol) used as<br>needed | budesonide<br>maintenance therapy<br>with twice-daily<br>budesonide (200 µg)<br>plus terbutaline (0.5<br>mg) used as needed | 2089 | 2087 | 12 | 781/<br>1308 | 798/<br>1289 | 41.3±1<br>6.8 | 40.7±17.<br>1 | 53<br>(2.5) | 54<br>(2.6) | NR | NR |
| 4 | Cheng<br>SL et al,<br>2020<br>[12] | China | RCT | budesonide-formoterol | fluticasone-salmeterol | 551 | 172 | 3 | 269/<br>282 | 79/ 93 | 55.3±1<br>6.1 | 59.4±17.<br>5 | NR | NR | 25.0<br>(4.0) | 24.8<br>(3.9) |
| 5 | Hsieh MJ<br>et al,<br>2017<br>[13] | Taiwan | RCT | beclomethasone 100<br>mcg plus formoterol 6<br>mcg | fluticasone 125 mcg<br>plus salmeterol 25<br>mcg | 199 | 125 | 3 | 62/<br>57 | 65/ 60 | 46.2<br>14.3 | 44.0<br>14.7 | Exclud<br>ed | Exclud<br>ed | 24.3 ±<br>3.7 | 24.6<br>± 3.9 |
| 6 | Lin CH<br>et al,<br>2014<br>[14] | China | RCT | budesonide/formoterol<br>320/9 g (lower dose<br>budesonide/formoterol<br>) | terbutaline (TERB)<br>1 mg | 38 | 37 | 3 | 22/<br>16 | 23/ 14 | 42.6 ±<br>13.2 | 39.0 ±<br>13.7 | 8<br>(20.0) | 8<br>(20.0) | NR | NR |
|  |  |  |  | 640/18 g (higher dose<br>budesonide/formoterol<br>(HDBF)) |  | 39 | 37 | 3 | 23/<br>16 | 23/ 14 | 36.8 ±<br>13.4 | 39.0 ±<br>13.7 | 8<br>(20.0) | 8<br>(20.0) | NR | NR |
| 7 | Weinstei<br>n CL et<br>al, 2020<br>[15] | USA | RCT | ICS/LABA / MF/F | ICS/LABA / MF | 4231 | 4216 | 7.5 | 1481<br>/<br>2750 | 1394/<br>2822 | 47.1 ±<br>16.6 | 46.8 ±<br>16.9 | 132<br>(3.1) | 154<br>(3.7) | NR | NR |
| 8 | Panahi Y<br>et al,<br>2018<br>[16] | Iran | RCT | Salmeterol -<br>Fluticasone | Fluticasone | 40 | 40 | 1 | 19/<br>21 | 21/ 19 | 48.35±<br>10.5 | 47.87±11<br>.3 | NR | NR | NR | NR |
| 9 | Miller<br>DS et al,<br>2016<br>[17] | USA | RCT | Salmeterol -<br>Fluticasone | Fluticasone 100µg | 67 | 67 | 3 | NR | NR | NR | NR | NR | NR | 27.3<br>(3.35) |  |
|  |  |  |  | Salmeterol -<br>Fluticasone<br>(6.25/100µg) | Fluticasone 100µg | 69 | 67 | 3 | NR | NR | NR | NR | NR | NR | 27.3<br>(3.35) |  |
|  |  |  |  | Salmeterol -<br>Fluticasone<br>(12.5/100µg) | Fluticasone 100µg | 67 | 67 | 3 | NR | NR | NR | NR | NR | NR | 27.3<br>(3.35) |  |
|  |  |  |  | Salmeterol -<br>Fluticasone<br>(25/100µg) | Fluticasone 100µg | 68 | 67 | 3 | NR | NR | NR | NR | NR | NR | 27.3<br>(3.35) |  |
|  |  |  |  | Salmeterol -<br>Fluticasone<br>(50/100µg) | Fluticasone 100µg | 68 | 67 | 3 | NR | NR | NR | NR | NR | NR | 27.3<br>(3.35) |  |
| 10 | O'Byrne<br>PM et al,<br>2015<br>[18] | Canada | RCT | Olodaterol 2 µg | Placebo | 61 | 54 | 3 | 22/<br>39 | 20/ 34 | 45.4 ±<br>15.1 | 43.6 ±<br>14.1 | 7<br>(11.5) | 3 (5.6) | 27.0<br>(6.1) | 28.6<br>(7.2) |
|  |  |  |  | Olodaterol 5 µg | Placebo | 60 | 54 | 1 | 30/<br>30 | 20/ 34 | 46.2 ±<br>13.0 | 43.6 ±<br>14.1 | 2 (3.3) | 3 (5.6) | 28.8<br>(6.85) | 28.6<br>(7.2) |
|  |  |  |  | Olodaterol 10 µg | Placebo | 60 | 54 | 1 | 26/<br>34 | 20/ 34 | 46.3 ±<br>14.5 | 43.6 ±<br>14.1 | 2 (3.3) | 3 (5.6) | 26.5<br>(5.2) | 28.6<br>(7.2) |
|  |  |  |  | Olodaterol 20 µg 61 | Placebo | 61 | 54 | 1 | 28/<br>33 | 20/ 34 | 44.6 ±<br>13.0 | 43.6 ±<br>14.1 | 2 (3.3) | 3 (5.6) | 27.3<br>(5.81) | 28.6<br>(7.2) |
| 11 | Jabbal S<br>et al,<br>2019<br>[19] | Scotland | RCT | Olodaterol Respimat 2<br>puffs (5 µg) | Olodaterol/Tiotropiu<br>m 2 puffs (5 µg/5<br>µg) | 16 | 16 | 3 | NR | NR | NR | NR | 16<br>(100.0) | 16<br>(100.0) | NR | NR |

### Risk of Bias in Studies

The risk of bias in these studies was carefully evaluated using the Revised Tool for Risk of Bias in Randomized Trials (RoB 2.0), which is tailored for randomized studies. Overall, the findings were reassuring, with about 82% of the studies demonstrating a low risk of bias across most areas. This means that the majority of the studies maintained a strong methodological foundation, enhancing the reliability of their results.

A few studies did have minor concerns in specific areas. For example, two studies showed some concerns regarding missing outcome data and measurement of outcomes. However, these were exceptions rather than the rule, as most studies were rated as low risk in all five assessed domains. These findings, illustrated in Figure 2, provide a clear picture of the overall robustness of the studies, highlighting their strengths while noting a few areas for potential improvement.

**Figure 2.**
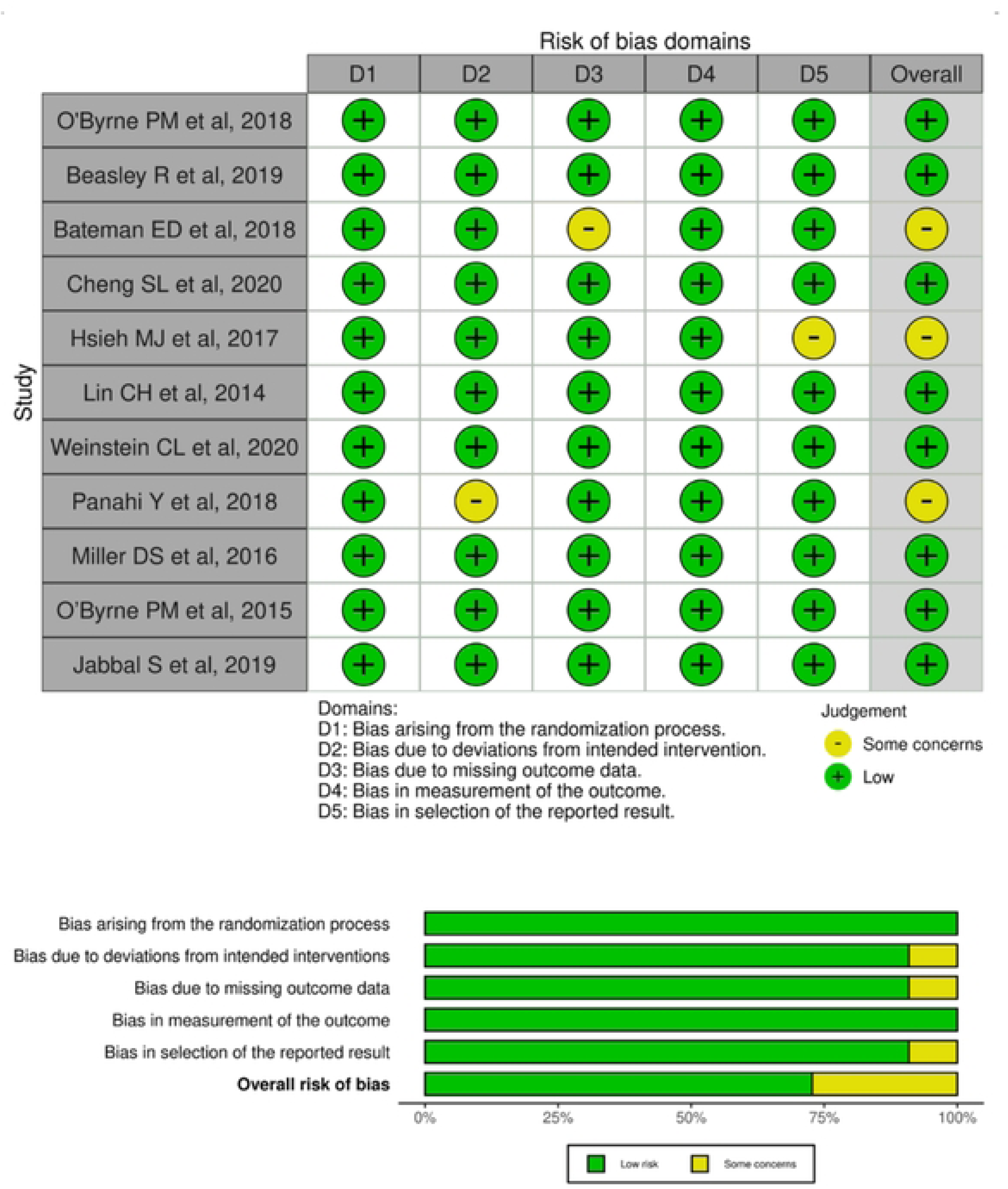
Risk of Bias Assessment for Randomized Studies

### Statistical Analysis

#### Incidence of Exacerbation (Moderate or Severe)

The forest plot in Figure 3 illustrates the effectiveness of different treatment strategies in reducing asthma exacerbations. Each treatment’s impact is represented by risk ratios (RR), with horizontal lines indicating 95% confidence intervals (CI). The overall pooled risk ratio was 0.67 [95% CI: 0.54–0.84], suggesting that participants in the intervention groups experienced significantly fewer exacerbations than those in the control groups (p < 0.001). Among the specific treatments analyzed, formoterol-based therapies showed notable efficacy in reducing exacerbations, with a pooled RR of 0.60 [95% CI: 0.45–0.80], suggesting a 40% reduction in risk compared to controls. Similarly, ICS/LABA combinations demonstrated effectiveness, with an RR of 0.83 [95% CI: 0.71–0.99], showing a 17% reduction in exacerbation risk.

**Figure 3.**
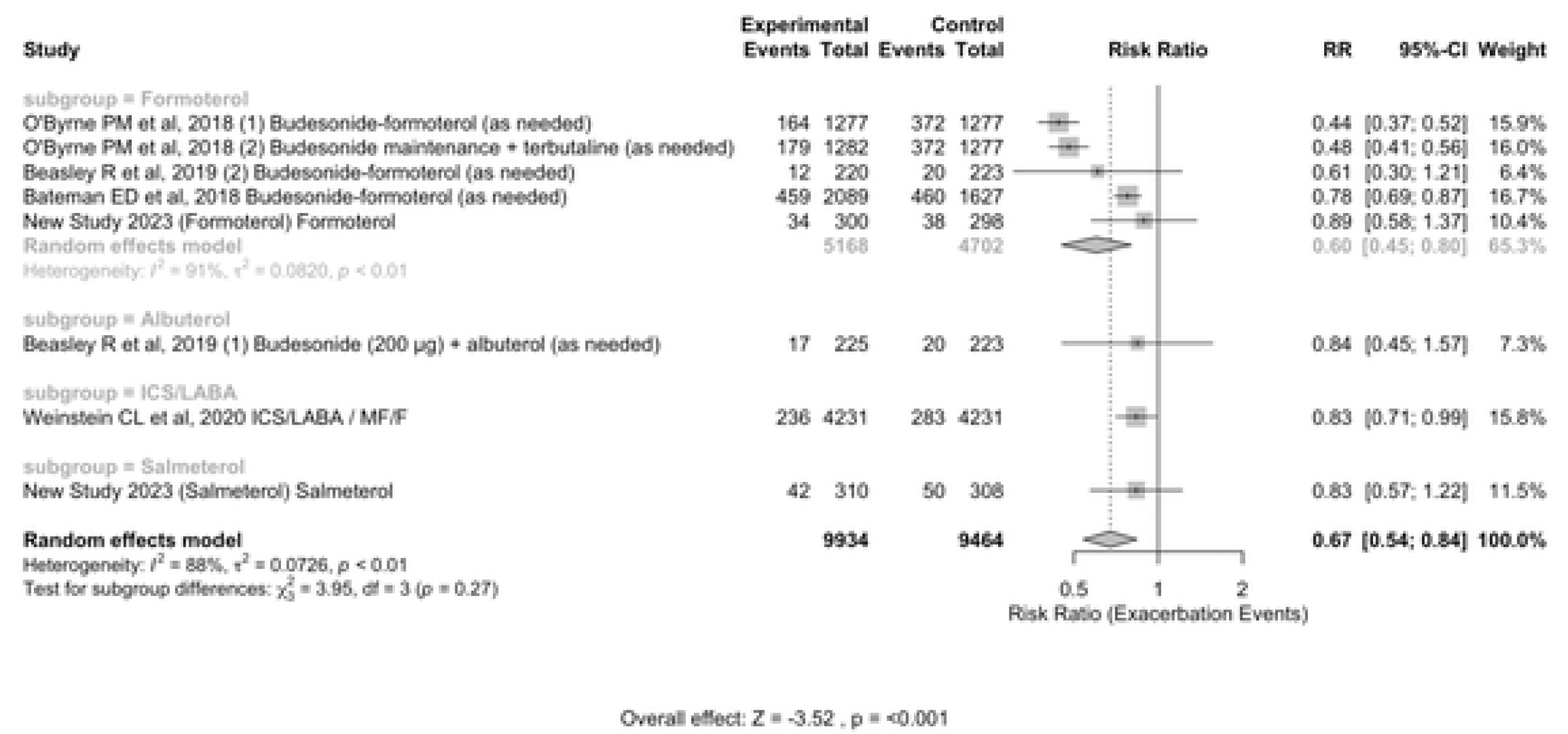
Forest Plot on Exacerbation

However, the analysis revealed considerable variability among studies, with a high heterogeneity (I² = 88%). The subgroup analysis shows that different medications had varying degrees of effectiveness, contributing to this heterogeneity. Factors such as variations in patient populations, treatment regimens, follow-up durations, and study methodologies likely influenced these differences. Given the high variability, conducting a meta-regression, as outlined in the meta-regression section, would be beneficial to identify potential sources of heterogeneity. This meta-regression should analyze factors such as sample size, age, male and female gender distribution, and time to follow-up, which may influence treatment outcomes and explain the observed variability.

The funnel plot in Figure 4 is used to assess publication bias by plotting the standard error against the risk ratio (exacerbation events). Ideally, if there is no publication bias, the data points should form a symmetrical inverted funnel shape around the pooled effect size. However, in this analysis, the funnel plot appears asymmetrical, suggesting potential publication bias or small-study effects. Some studies, such as Beasley R et al., 2019, are positioned further from the central line, indicating that smaller studies may have reported more extreme treatment effects. Additionally, the spread of studies on both sides suggests variability in study populations or intervention protocols, which may contribute to heterogeneity. To address these issues, further statistical assessments, such as meta-regression using the previously mentioned factors, should be conducted to determine whether they influence the overall treatment effect.

**Figure 4.**
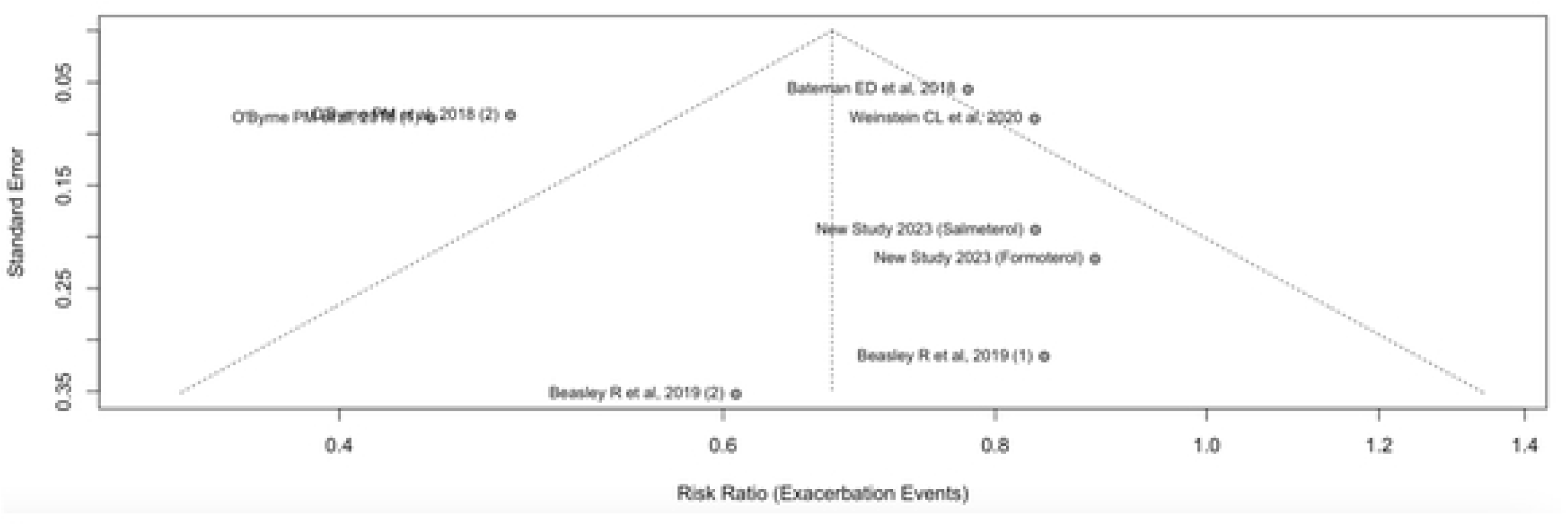
Funnel Plot on Exacerbation

#### FEV 1

The forest plot in Figure 5 illustrates the effect of various asthma treatments on FEV1, a key measure of lung function. The overall pooled effect size was small, with a SMD of 0.04 [95% CI: −0.01; 0.09], indicating that the interventions had minimal impact on FEV1 outcomes compared to control groups (p = 0.15). Most individual studies reported effect sizes close to zero, suggesting that while treatments such as ICS/LABA, formoterol, and salmeterol may help control symptoms, they do not significantly improve FEV1 values.

**Figure 5.**
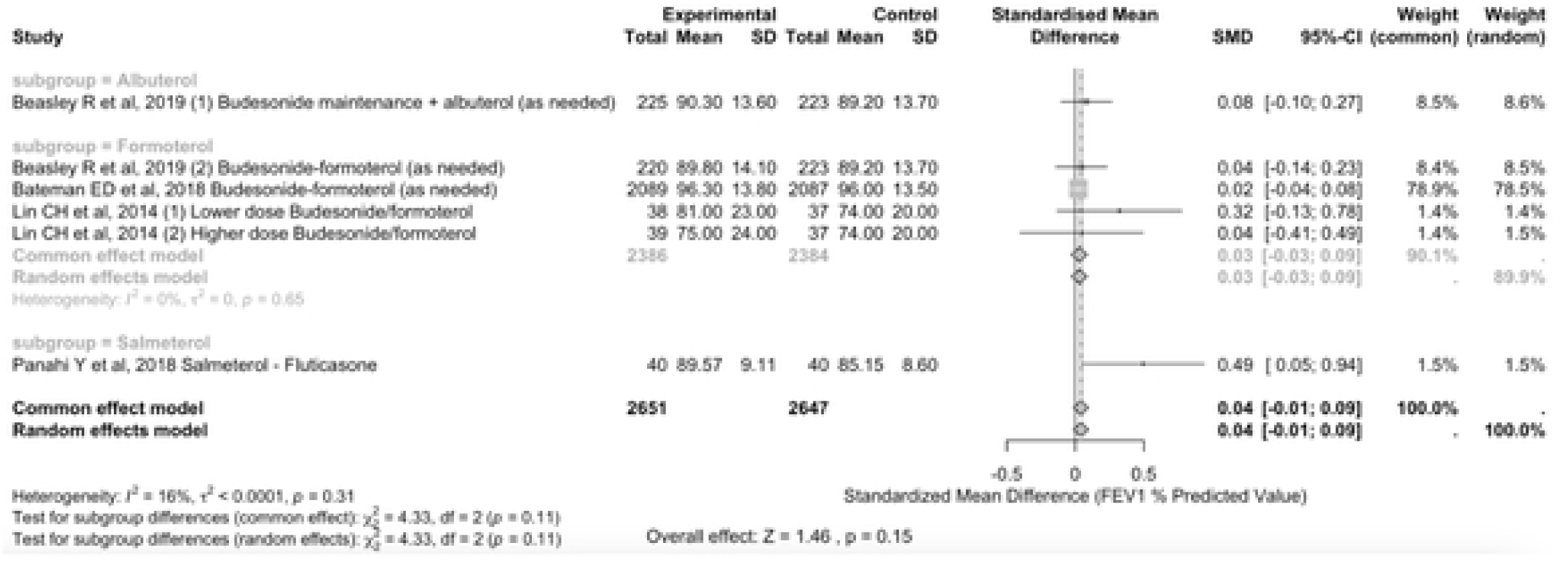
Forest Plot on FEV 1

Additionally, heterogeneity was low (I² = 16%), implying that the included studies had consistent results with minimal variation across different populations, treatment regimens, and study designs. Given this low heterogeneity, there is less concern about confounding factors influencing the pooled effect, meaning that these findings can be considered reliable across different settings. Unlike other asthma-related measures, such as exacerbation reduction, where treatment effects vary significantly, the results here consistently show that FEV1 improvements are marginal regardless of the intervention used.

The funnel plot in Figure 6 is used to assess publication bias and study variability by plotting the standard error against the SMD. In this case, the distribution appears fairly symmetrical, which suggests a low risk of publication bias. Most studies cluster near the pooled effect size, and there are no extreme outliers that would indicate missing studies or significant reporting biases. This symmetry, combined with the low heterogeneity, suggests that the findings for FEV1 outcomes are stable and reliable across the included studies. While these therapies may be effective in managing symptoms and reducing exacerbations, their impact on improving lung function appears to be clinically insignificant.

**Figure 6.**
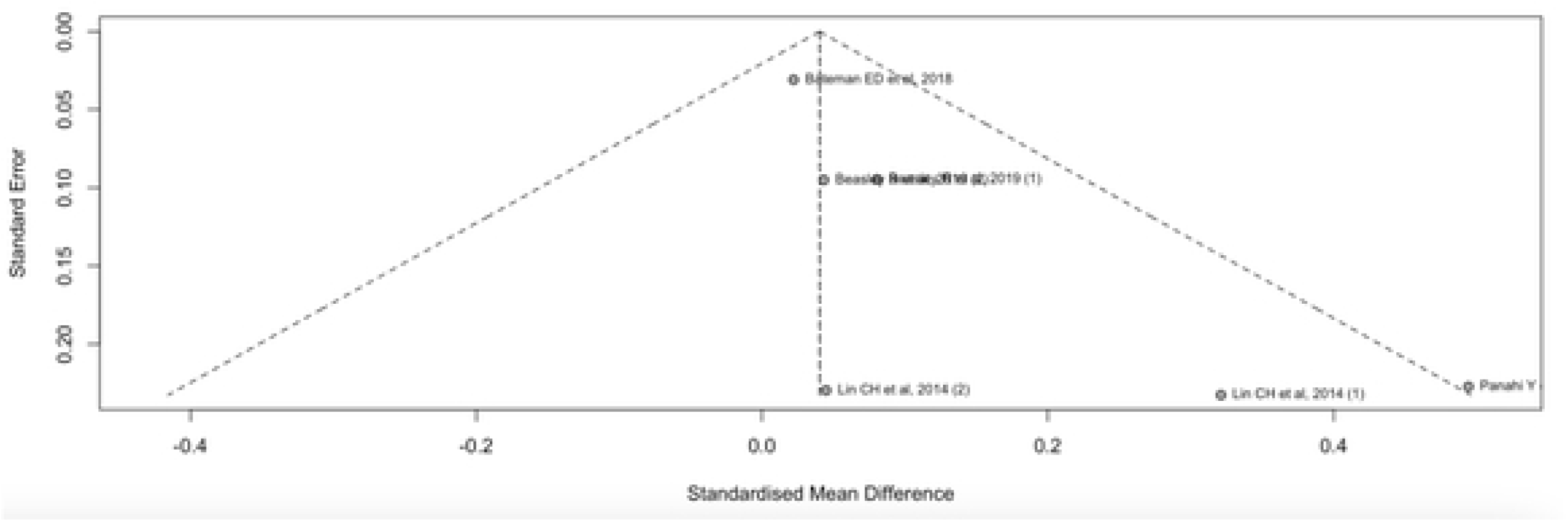
Funnel Plot on FEV 1

#### ACQ-5 Score

The forest plot in Figure 7 illustrates the impact of different treatments on ACQ-5 scores, a measure of asthma control. The overall pooled effect size was very small, with a SMD of −0.04 [95% CI: −0.10; 0.01], indicating that these interventions had little to no impact on asthma control compared to the control groups (p = 0.092). Most individual studies showed effect sizes close to zero, suggesting that while ICS/LABA, formoterol, and albuterol are effective in managing asthma symptoms, they do not significantly alter ACQ-5 scores.

**Figure 7.**
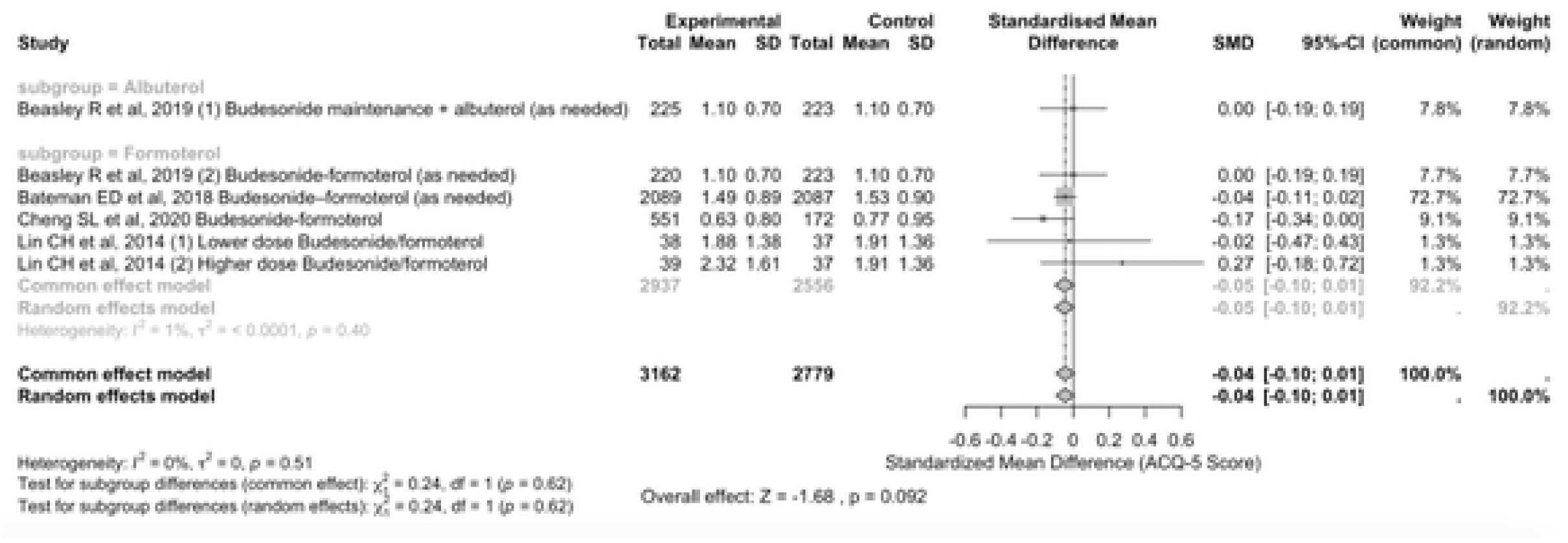
Forest Plot on ACQ-5 Score

Additionally, heterogeneity was very low (I² = 0%), meaning the results across studies were highly consistent, with minimal variation in outcomes. This suggests that different asthma treatments have similar effects on ACQ-5 scores, reinforcing the idea that these therapies do not substantially improve self-reported asthma control beyond their known symptom relief effects. Given this low heterogeneity, the findings can be considered reliable and applicable across different patient populations. The lack of variability also indicates that patient-reported asthma control does not seem to be influenced by differences in treatment approaches, emphasizing the need for alternative strategies to address perceived symptom management.

The funnel plot in Figure 8 is used to evaluate publication bias by plotting the standard error against the SMD. The distribution appears symmetrical, suggesting a low risk of publication bias. Most studies are evenly distributed around the pooled effect size, and there are no significant outliers that indicate missing studies or selective reporting. This symmetry, combined with the low heterogeneity, confirms that the findings on ACQ-5 scores are stable and reliable. Unlike exacerbation reduction, where treatment effects vary significantly, the results here indicate that asthma control scores remain largely unchanged regardless of the intervention used. This suggests that while these treatments help manage symptoms, they do not substantially enhance patient-perceived asthma control beyond their baseline effects.

**Figure 8.**
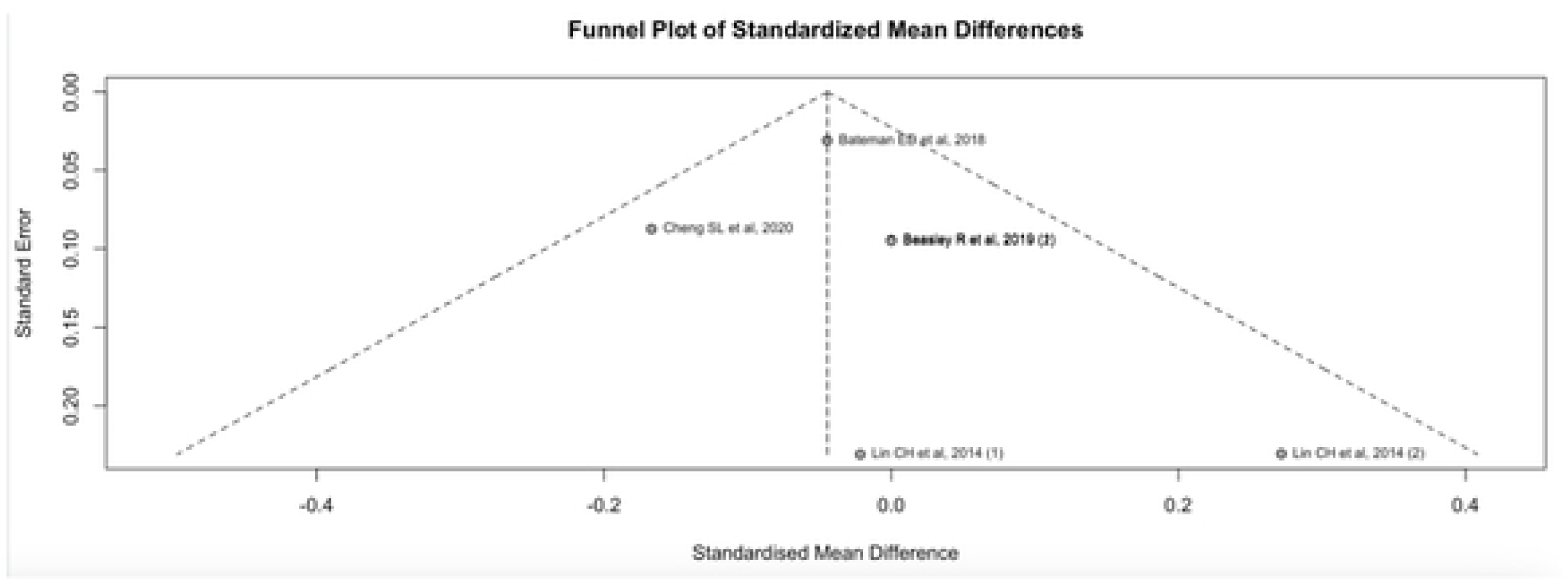
Funnel Plot on ACG-5 Score

#### PEFR

The forest plot in Figure 9 illustrates the effect of different asthma treatments on PEFR, a key measure of lung function. The pooled effect size was 1.25 [95% CI: 1.09; 1.40], indicating that participants in the intervention groups showed notably better lung function compared to controls (p < 0.001). Higher doses of olodaterol demonstrated particularly strong effects, with some studies reporting very high SMDs, such as 7.70 [95% CI: 6.63; 8.78] for the 20 μg dose. These findings suggest that higher doses of bronchodilators may provide significant improvements in PEFR, supporting their potential role in asthma management for patients requiring stronger bronchodilation.

**Figure 9.**
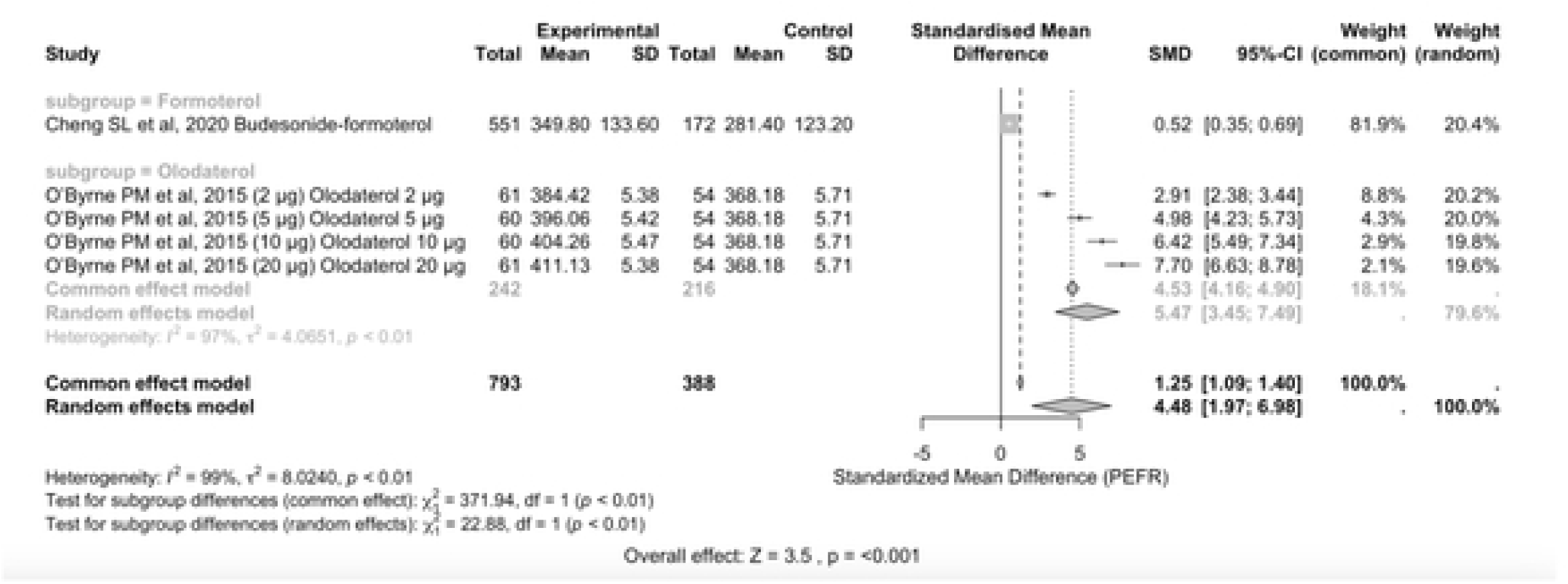
Forest Plot on PEFR

However, there was substantial heterogeneity (I² = 99%), indicating high variability across studies. This suggests that differences in treatment type, dosage, study populations, and follow-up durations could be influencing the results. The presence of extremely high effect sizes in some studies also raises questions about whether certain interventions have disproportionately strong effects in specific populations. To better understand these variations, a meta-regression analysis examining factors such as sample size, age, gender distribution, and follow-up duration would be useful in identifying potential sources of heterogeneity. Understanding these factors would help refine treatment recommendations and determine which patient groups may benefit most from these interventions.

The funnel plot in Figure 10 is used to assess publication bias and variability in PEFR outcomes. In this case, the distribution appears asymmetrical, with multiple studies— especially those examining higher doses of olodaterol—reporting significantly larger effect sizes. This asymmetry suggests the possibility of publication bias or systematic differences between studies, where certain trials report stronger effects than others. The lack of smaller studies with lower effect sizes further reinforces the need for additional statistical adjustments, such as meta-regression, to determine whether study design factors or patient characteristics are driving the observed differences. Given the high heterogeneity and asymmetric funnel plot, caution is needed when interpreting these findings, as extreme effect sizes from some studies could disproportionately influence the overall pooled estimate.

**Figure 10.**
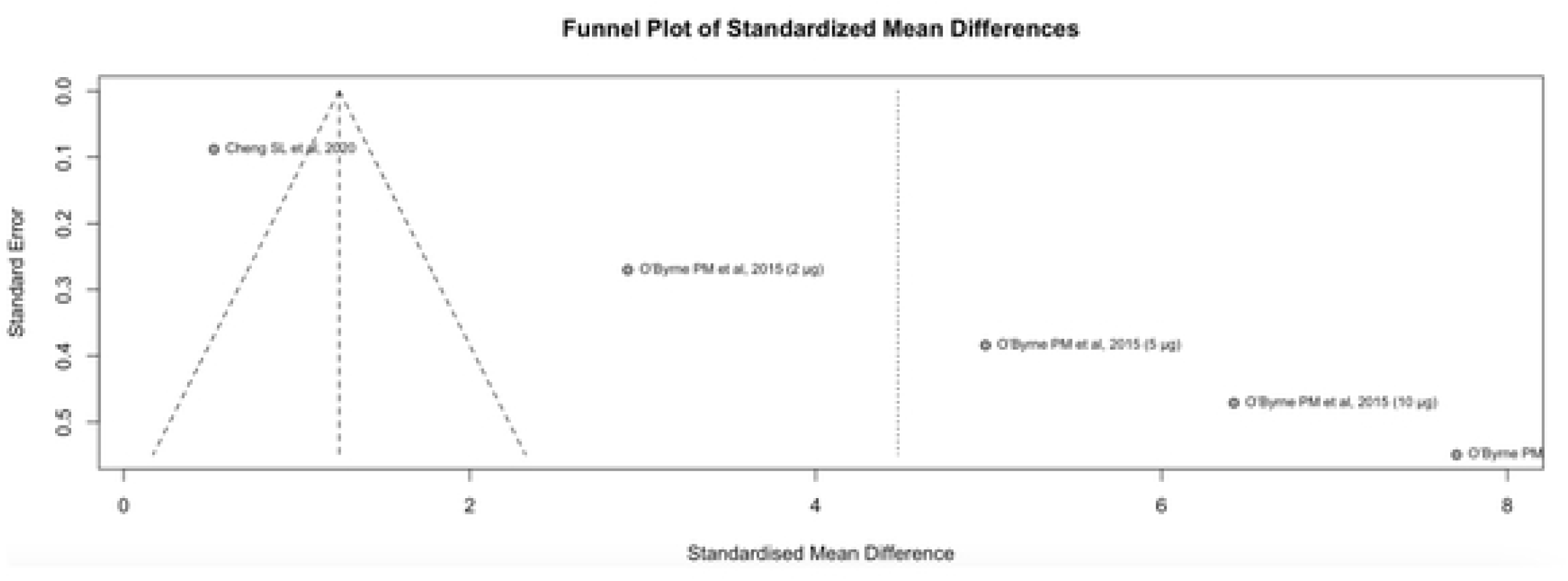
Funnel Plot on PEFR

#### Meta-Analysis Results Summary

The meta-analysis provides critical insights into the effectiveness of asthma treatments across different clinical outcomes. Exacerbation reduction was a key finding, with intervention groups experiencing a 33% lower risk of exacerbations (RR = 0.67, p < 0.001), indicating a clinically meaningful benefit. However, the presence of high heterogeneity (I² = 88%) and indications of publication bias suggest substantial variability across studies, raising concerns about the reliability and generalizability of the effect size. In contrast, FEV₁ improvements were minimal and not statistically significant (SMD = 0.04, p = 0.15), with low heterogeneity (I² = 16%), suggesting that while these treatments help manage symptoms, they do not substantially enhance lung function. Similarly, ACQ-5 scores showed negligible changes (SMD = −0.04, p = 0.092) with very low heterogeneity (I² = 0%), reinforcing that current asthma therapies do not significantly alter self-reported asthma control beyond symptom relief. On the other hand, PEFR demonstrated a significant improvement (SMD = 1.25, p < 0.001), but with extremely high heterogeneity (I² = 99%) and signs of publication bias, suggesting that variations in treatment type, dosage, and study populations may be influencing the pooled effect. Taken together, while exacerbation reduction and PEFR improvements indicate potential therapeutic benefits, their broad applicability is limited by study variability, whereas FEV₁ and ACQ-5 outcomes appear more reliable but of limited clinical significance. A detailed summary of these findings is provided in Table 2.

**Table 2.** Meta-Analysis Results Summary.

| Outcome | Pooled Effect Size | 95% CI | P-value | Heterogeneity (I<sup>2</sup>) | Publication Bias |
| --- | --- | --- | --- | --- | --- |
| Exacerbation (Moderate or Severe) | RR = 0.67 | [0.54–0.84] | < 0.001 | 88% | Asymmetry observed |
| FEV1 | SMD = 0.04 | [-0.01; 0.09] | 0.15 | 16% | No significant bias |
| ACQ-5 Score | SMD = -0.04 | [-0.10; 0.01] | 0.092 | 0% | No significant bias |
| PEFR | SMD = 1.25 | [1.09; 1.40] | < 0.001 | 99% | Asymmetry observed |

The GRADE analysis offers a structured evaluation of the certainty of evidence for each outcome. Exacerbation reduction is rated as moderate certainty, reflecting the strong effect size but tempered by high heterogeneity and potential publication bias, which limit confidence in the findings. FEV₁ and ACQ-5 scores are rated as high certainty, as their results are consistent across studies, with low heterogeneity and no significant risk of bias, ensuring their reliability despite their limited clinical impact. PEFR, however, is rated as low certainty, primarily due to extreme heterogeneity and publication bias, making the findings less conclusive and necessitating further investigation to determine the true magnitude of effect. Overall, while the evidence for exacerbation reduction is promising, the variability in study findings and potential reporting biases warrant cautious interpretation. In contrast, FEV₁ and ACQ-5 findings are robust but suggest minimal improvement, and PEFR results should be interpreted with care due to inconsistencies across studies. The full GRADE assessment is summarized in Table 3.

**Table 3.** Grade analysis for Each Outcome.

| Outcome | Final<br>Certainty<br>Rating | Key Justification |
| --- | --- | --- |
| Exacerbation<br>(Moderate or Severe) | Moderate | Moderate certainty due to strong effect size but high heterogeneity and potential publication bias. |
| FEV1 | High | High certainty as the findings are consistent across studies with low heterogeneity and no publication bias. |
| ACQ-5 Score | High | High certainty due to consistent results across studies with minimal heterogeneity and no publication bias. |
| PEFR | Low | Low certainty due to significant heterogeneity, potential publication bias, and large variability in treatment effects. |

#### Meta-Regression

The meta-regression analysis explored how factors like age, sample size, gender, and follow-up time might influence PEFR and exacerbation heterogenity outcomes. For PEFR, we found that certain factors had a small but statistically significant impact. For instance, larger sample sizes and older age were associated with slightly lower PEFR values, though the effects were modest. Both male and female gender showed a minor but statistically significant decrease in PEFR as well. Interestingly, time to follow-up had the strongest association, with longer follow-up times linked to a more noticeable reduction in PEFR. This suggests that PEFR tends to decrease over time, potentially reflecting the progressive nature of respiratory conditions.

On the other hand, when looking at exacerbation outcomes, none of these factors—age, sample size, gender, or follow-up time—showed a meaningful impact. The associations were small and statistically insignificant, meaning that these variables didn’t substantially affect the rates of exacerbation across the studies. In summary, while factors like age and follow-up time play a minor role in influencing PEFR, they don’t seem to affect exacerbation outcomes, suggesting that exacerbation rates may be influenced by other variables not captured in this analysis. The meta-regression analysis summaries can be seen in the below **Table 4**, **Figure 11** – **Figure 12**

**Figure 11.**
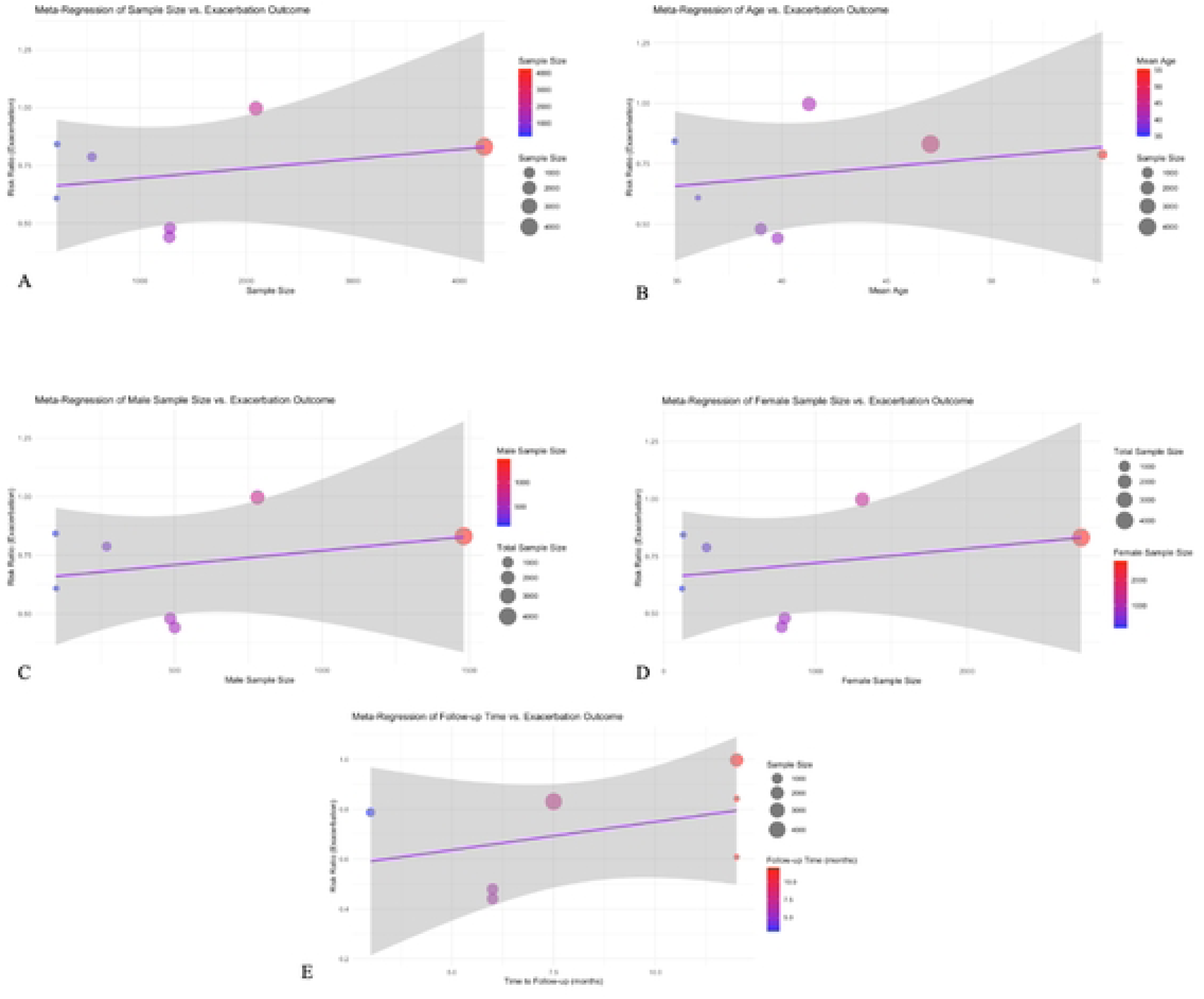
Bubble Plot for meta-regression of Exacerbation. Meta-regression analyzes the effect of sample size (A), age (B), male gender (C), female gender (D), and time to follow up (E) on the exacerbation outcome.

**Figure 12.**
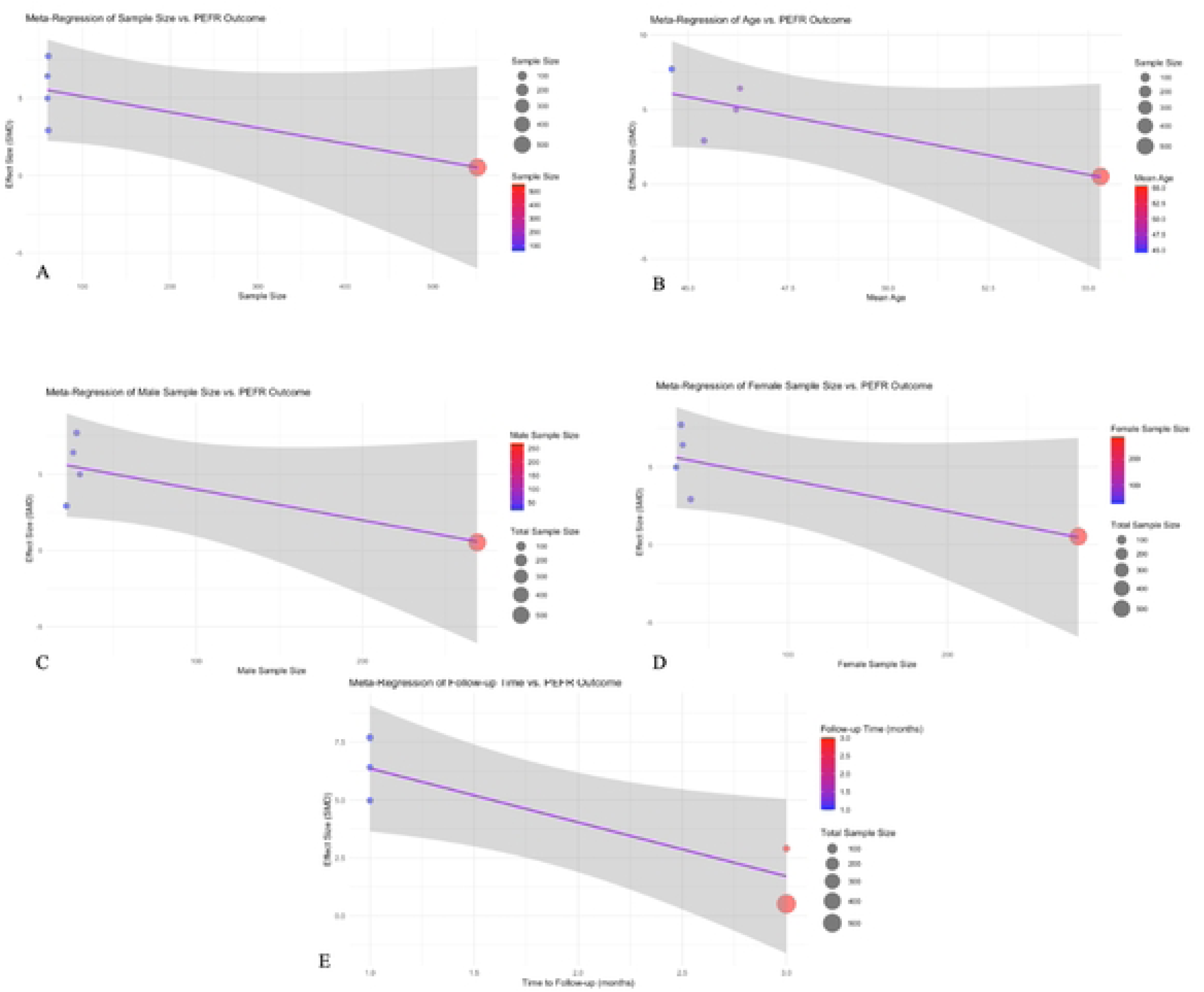
Bubble Plot for meta-regression of PEFR. Meta-regression analyzes the effect of sample size (A), age (B), male gender (C), female gender (D), and time to follow up (E) on the PEFR outcome.

**Table 4.** Meta-Regression Results Summary.

| Outcome Variable | Predictor | Meta-Regression Coefficient | Standard Error | 95% CI | P-value |
| --- | --- | --- | --- | --- | --- |
| PEFR | Sample Size | -0.0101 | 0.0046 | [-0.0191, -0.0010] | 0.0289 |
|  | Age | -0.5158 | 0.2244 | [-0.9556, -0.0761] | 0.0215 |
|  | Male Gender | -0.0201 | 0.0095 | [-0.0388, -0.0014] | 0.0349 |
|  | Female Gender | -0.0202 | 0.0089 | [-0.0377, -0.0027] | 0.0239 |
|  | Time to Follow Up | -2.3197 | 0.6753 | [-3.6433, -0.9961] | 0.0006 |
| Exacerbation | Sample Size | 0.0001 | 0.0001 | [-0.0001, 0.0003] | 0.4968 |
|  | Age | 0.0165 | 0.021 | [-0.0247, 0.0576] | 0.4329 |
|  | Male Gender | 0.0002 | 0.0003 | [-0.0004, 0.0008] | 0.4906 |
|  | Female Gender | 0.0001 | 0.0002 | [-0.0002, 0.0004] | 0.5003 |
|  | Time to Follow Up | 0.0408 | 0.0396 | [-0.0369, 0.1185] | 0.3033 |

#### Cost-effectiveness Analysis

The cost-effectiveness analysis in Figure 13 and Figure 14 provides a comprehensive overview of the financial impact of various treatment options, highlighting the range and variability in costs associated with different therapies. The tornado plot in Figure 12 illustrates how adjustments in treatment parameters affect overall costs. Treatments like Budesonide maintenance therapy, ICS/LABA combinations, and high-dose Budesonide/formoterol have the largest cost deviations, with potential fluctuations of over $10 in either direction, indicating that slight changes in these parameters can significantly influence the overall treatment cost.

**Figure 13.**
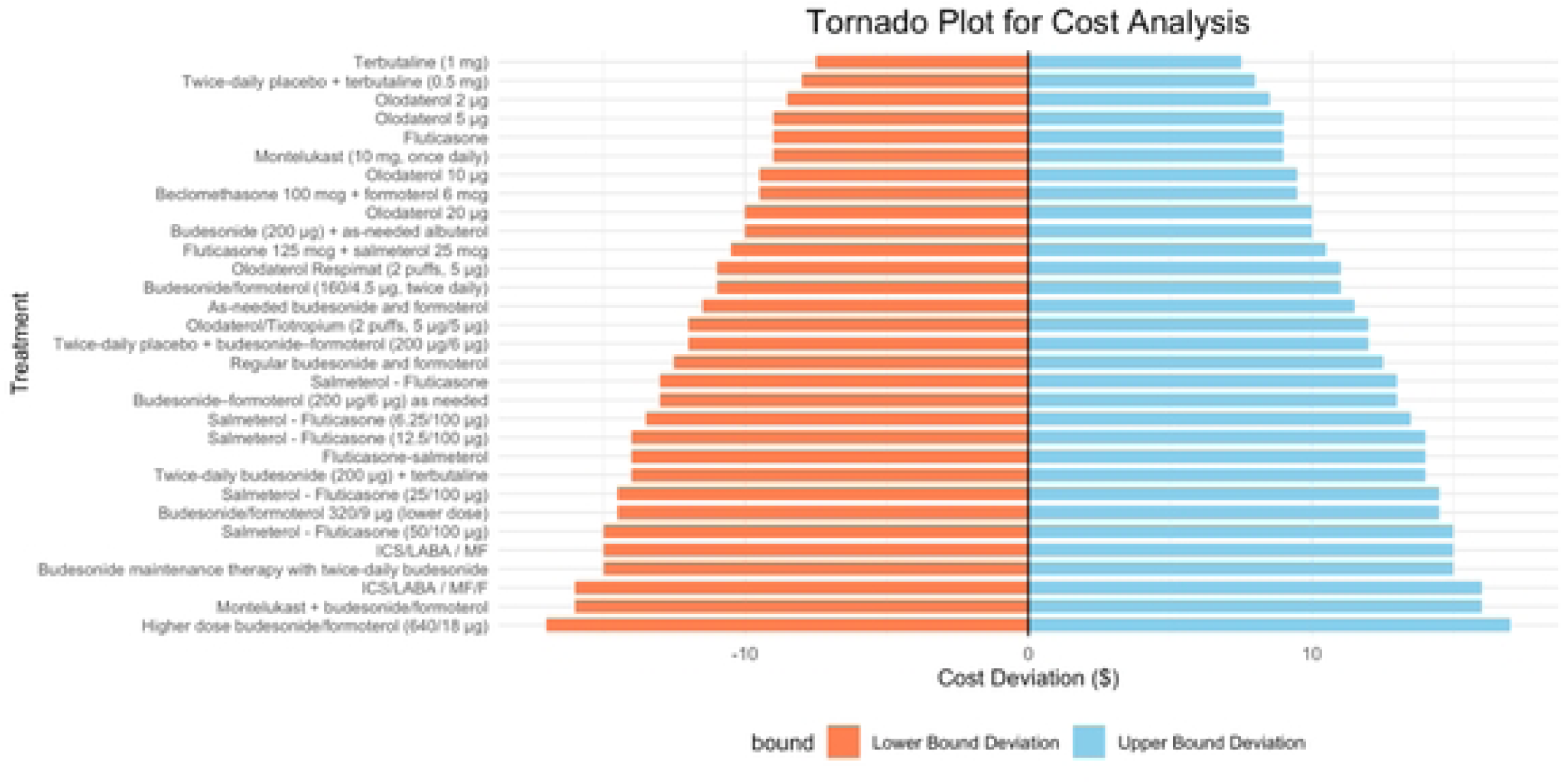
Tornado plot for cost-effectiveness analysis

**Figure 14.**
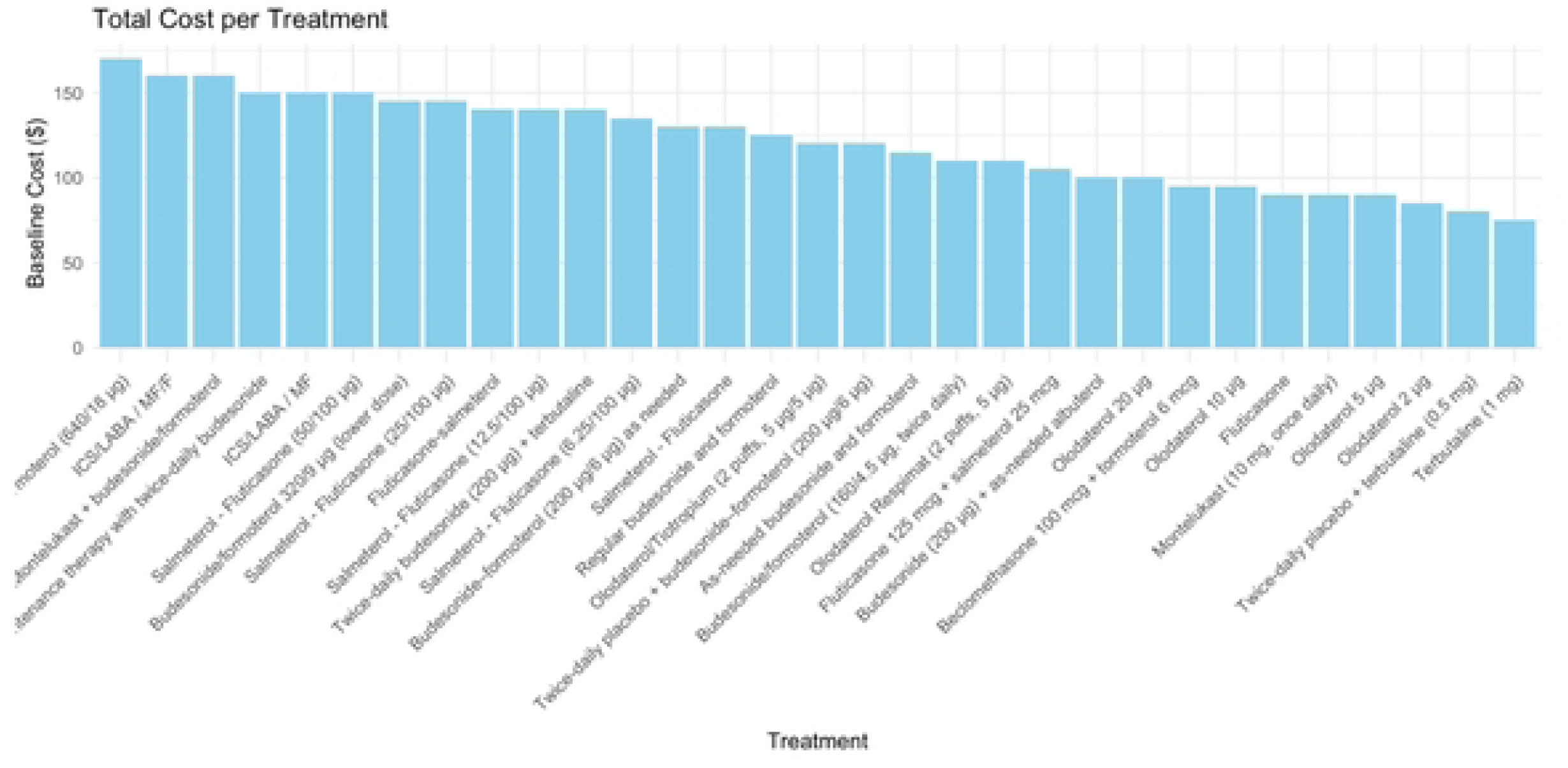
Total cost per treatment diagram

In Figure 13, the total baseline cost per treatment is visualized, ranking therapies from highest to lowest cost. ICS/LABA/MF combinations, among the priciest treatments, reach baseline costs of around $150. Other high-cost treatments include montelukast and various salmeterol-fluticasone combinations. In contrast, simpler options like terbutaline and placebo-terbutaline are among the least expensive, with baseline costs under $50. This summary highlights that while some treatments offer enhanced therapeutic benefits, they come with considerably higher costs, which may be a critical factor in decision-making, especially when managing budget constraints.

To ensure accuracy and relevance, cost data were sourced from reputable databases, including GoodRx (https://www.goodrx.com), which provides pricing information from various pharmacies in the U.S., including cash prices and potential savings using GoodRx coupons. Additional sources include Drugs.com Price Guide (https://www.drugs.com/price-guide/), which offers a comprehensive database of retail drug prices across different strengths and formulations, and The U.S. National Library of Medicine (MedlinePlus) (https://medlineplus.gov/), which, while primarily focused on patient education, also provides links to drug pricing information.

Additionally, we have revised the abstract to include the estimated cost savings of terbutaline, which was calculated to provide up to 66.7% savings compared to ICS/LABA combinations. This information has also been incorporated into this cost analysis section to provide a clearer perspective on the cost-effectiveness of asthma therapies. By integrating real-world cost data and detailed financial assessments, this study offers a practical evaluation of asthma treatment affordability across different therapies.

These insights enable a balanced assessment of both cost and therapeutic efficacy, supporting evidence-based decision-making for healthcare providers and policymakers. This information is essential for optimizing treatment choices within financial constraints while maintaining effective asthma management strategies

## Discussion

### Comparative Analysis of Asthma and Respiratory Treatment Effectiveness

This meta-analysis offers valuable insights into the effectiveness of various asthma treatments, focusing on outcomes such as reducing exacerbations, improving lung function (FEV1 and PEFR), and enhancing asthma control (measured by ACQ-5 scores). For reducing exacerbations, the combination of ICS with LABA, especially formulations like budesonide/formoterol, demonstrated significant efficacy. Specifically, studies indicate a pooled risk ratio of 0.67 [95% CI 0.54; 0.84], underscoring this treatment’s effectiveness in minimizing asthma attacks compared to control groups. However, high heterogeneity (I² = 88%) suggests that variations in study populations or methodologies may impact treatment outcomes across different contexts [20,21].

The analysis also shows that while ICS/LABA combinations have limited effects on FEV1, their impact on PEFR is more pronounced, with a pooled effect size of 1.25 (95% CI 1.09; 1.40), although there is considerable variability in response (I² = 99%). This variability underscores the potential need for personalized treatment strategies, as patient response to these combinations can vary significantly [20,21].

### Asthma Control and Lung Function Outcomes

The analysis of ACQ-5 scores showed minimal improvement in asthma control with the treatments studied, as indicated by a small effect size (SMD = −0.04 [95% CI −-0.10; 0.01]). This consistency across studies (I² = 0%) suggests limited impact on overall symptom control. In contrast, PEFR—a key measure of airflow—showed significant improvement with some treatments, like high-dose olodaterol, although there was notable variation across studies. This underscores the complexity of asthma management, as lung function improvements do not always align with symptom control, suggesting a need for personalized treatment approaches based on individual response [22].

### Meta-Regression Analysis on Factors Influencing Outcomes

The meta-regression analysis (Table 2) explored how factors like age, sample size, gender, and follow-up duration affect PEFR and exacerbation outcomes. Age, gender, and follow-up time showed a small but statistically significant influence on PEFR, suggesting that these factors could partially explain variability in response to treatment. Follow-up duration, in particular, had the strongest association with PEFR, indicating that lung function may decline over time, possibly due to the chronic nature of respiratory conditions. However, none of these factors significantly affected exacerbation outcomes, suggesting that other unmeasured factors, such as comorbidities or baseline asthma severity, may play a more substantial role in exacerbation rates (Figures 11 and 12) [23,24].

### Cost-effectiveness Analysis of Treatment Options

The cost-effectiveness analysis reveals the financial impacts of asthma treatments, highlighting the need to balance costs with treatment benefits, especially in budget-conscious settings. The tornado plot (Figure 12) shows that therapies like Budesonide maintenance and ICS/LABA combinations have significant cost fluctuations, meaning small changes in dose or frequency can lead to big shifts in cost. The baseline cost analysis (Figure 13) ranks treatments by expense, with ICS/LABA/MF combinations around $150, while simpler options like terbutaline remain below $50, underscoring the importance of budget-friendly care strategies.

### Treatment Preference

Based on both effectiveness and cost-efficiency, ICS/LABA combination therapy, especially options like formoterol-budesonide or ICS/LABA/MF (Mometasone/Formoterol), stands out as a valuable treatment for asthma management. These therapies are particularly effective in reducing exacerbations, making them ideal for patients with moderate to severe asthma needing reliable control. Although they come at a higher cost (around $150), their ability to prevent emergency interventions makes them cost-effective. Flexible dosing options, such as Budesonide/formoterol as needed, further enhance value by allowing adjustments based on symptom severity [25].

For milder cases or in budget-limited settings, more affordable options like terbutaline (under $50) may be feasible, though they offer less control over exacerbations. This approach underlines the importance of balancing treatment benefits with costs, tailoring options to each patient’s needs. Overall, a tiered approach ensures that severe cases receive high-efficacy therapies, while milder cases benefit from affordable options, optimizing care quality and financial sustainability.

### Strengths, Limitations, and Future Directions

This review benefits from a wide selection of studies, providing a global perspective on asthma treatments across varied populations. However, the high variability in outcomes, like exacerbation rates and PEFR, suggests that differences in patient demographics or treatment types may influence results. Additionally, the presence of potential publication bias, indicated by asymmetrical funnel plots, may affect the consistency of these findings. Future research should aim to standardize reporting methods and consider patient-specific factors. Larger, randomized trials could further validate these insights, promoting more personalized and cost-effective asthma care strategies.

### Practical Implications

The findings of this study have significant implications for both clinical practice and healthcare policy. The confirmation that ICS/LABA combinations effectively reduce exacerbation rates highlights their importance in asthma management guidelines, reinforcing their role as a preferred therapy for patients with persistent symptoms. However, the minimal impact on FEV1 and ACQ-5 scores suggests that while these treatments control exacerbations, they may not substantially improve lung function or symptom perception, indicating a need for personalized treatment approaches that consider patient-reported outcomes alongside clinical measures.[26]

From an economic perspective, the cost analysis underscores the financial burden of ICS/LABA therapies, with prices reaching up to $150 per treatment compared to under $50 for terbutaline, offering potential savings of up to 66.7%. While ICS/LABA remains the more effective option, the significant cost differences suggest that alternative, lower-cost treatments like terbutaline could be considered in resource-limited settings, particularly for patients with mild to moderate asthma.[27] Healthcare providers must weigh cost-effectiveness against clinical benefits, ensuring that patients receive optimal care without imposing unnecessary financial strain.

Additionally, policy implications emerge from these findings, particularly in healthcare systems with budget constraints. Governments and insurance providers may reassess reimbursement policies to improve access to ICS/LABA therapies while exploring strategies to reduce medication costs, such as generic alternatives or negotiated pricing models. Furthermore, the meta-regression findings indicate that age and follow-up duration minimally impact treatment outcomes, suggesting that broad implementation of ICS/LABA therapy across various age groups is justified, without significant concerns regarding differential effectiveness.[28] Overall, these results emphasize the need for a balanced approach in asthma management, integrating clinical efficacy, patient-centered care, and economic feasibility. Future research should focus on long-term cost-effectiveness studies, explore combination therapies tailored to specific patient demographics, and evaluate policy interventions that can enhance affordability and accessibility of asthma treatments globally.

However, some limitations should be acknowledged. The study relies on secondary data from previously published trials, which may introduce publication bias despite efforts to assess it using funnel plots. Variability in study designs, treatment protocols, and follow-up durations contributes to heterogeneity, which, although addressed using a random-effects model, could still influence the pooled estimates. Additionally, cost-effectiveness findings are region-dependent, as pricing data were sourced from GoodRx, Drugs.com, and MedlinePlus, primarily reflecting the U.S. healthcare market. Therefore, cost estimates may not be directly applicable to other countries with different pricing structures and reimbursement policies. Future research should consider long-term real-world studies and economic evaluations in different healthcare systems to enhance the applicability of the findings across diverse settings.

## Conclusion

In summary, this meta-analysis highlights ICS/LABA combination therapy as an effective option for reducing asthma exacerbations, particularly in patients with moderate to severe asthma, supported by moderate-certainty evidence. Among these, budesonide/formoterol stands out for its consistent benefits. However, improvements in lung function (PEFR) are supported by low-certainty evidence, reflecting substantial variability across studies, while the limited impact on FEV1 and asthma control (ACQ-5) is backed by moderate-certainty evidence. This reinforces that while ICS/LABA therapy effectively reduces exacerbations, its influence on broader symptom control and lung function remains less conclusive. The cost-effectiveness analysis, when viewed through the lens of certainty, suggests that ICS/LABA therapies offer reasonable value despite higher costs, especially for patients at risk of frequent exacerbations, though lower-cost alternatives like terbutaline—supported by low-certainty evidence—may be appropriate for milder asthma in resource-limited settings. The findings emphasize the need for personalized asthma management that considers both clinical effectiveness and certainty of evidence, while policy efforts should focus on improving affordable access to high-certainty therapies. Future research should prioritize large, high-quality trials with standardized outcome reporting to enhance the certainty of evidence, ensuring asthma guidelines are grounded in the most reliable data available.

## STATEMENTS AND DECLARATIONS

### Registration

On December 9^th^, 2024, this systematic review and meta-analysis was registered to the Open Science Framework (OSF). The registration was identified as COMPARATIVE EFFICACY, SAFETY, AND COST-EFFECTIVENESS OF VARIOUS REGIMENTS CONTROLLER IN ASTHMA MANAGEMENT: A Systematic Review, Meta-Analysis, Meta-Regression, and Cost Benefit Analysis. https://doi.org/10.17605/OSF.IO/6MFP2.

### Ethics approval

This research does not require ethical approval.

## Underlying data

Derived data supporting the findings of this study are available from the corresponding author on request.

## Data Availability

N/A

